# An intermediate step in bridging the gap between evidence and practice: developing and applying a methodology for “general good practices”

**DOI:** 10.1101/2022.04.27.22274383

**Authors:** Heléna Safadi, Judit Lám, Ivett Baranyi, Éva Belicza

## Abstract

The gap between evidence and clinical practice has been in the focus of researches for decades. Although successful implementation means the new knowledge must work in particular environments, it doesn’t mean that the entire process should exclusively be executed by the individual institutes. This is the point where we assumed that an intermediate step, the “general good practice”, could help to ensure that translation is done in a more professional way.

The development of the general good practice methodology was based on our infinitE model, which organized the factors of successful translation into an evidence-editing-embedding-effect on practice framework, using tools from the disciplines of Evidence-Based Medicine, Quality Improvement and Change Management.

The methodology organised the editing and embedding part of the development into a process involving three full-day sessions carried out with different health professionals, experts and moderators. After pilot testing, it was finalized and applied to other topics as well.

The methodology presented in detail in this paper, centred on flow chart, process analysis, failure mode identification and Kotter’s 8-step model. Beside the pilot topic of the institutional process of resuscitation, the methodology has also proved applicable to more than ten other topics, meaning that at least all the core elements of the proposed bundle of general good practice have been produced in the development process.

Compared to the guidelines, general good practices demonstrate the evidence in operation, helping to develop workflows, responsibilities, documentation, trainings, etc. and can also be a starting point for the digitalisation of care processes.

The next step is to examine how healthcare institutions can build on these in their own editing and embedding activities, and what the results will be. Further studies could explore the applicability of the development methodology in different healthcare systems or at different levels of maturity in terms of quality.

## Introduction

The gap between evidence and daily clinical practice is widely known and has been in the focus of researches for decades. Investigating this problem and the underlying causes usually starts with identifying the barriers and facilitators to implementation.[1-9] In a scoping review, Fisher et al. grouped the barriers into three levels: personal factors that relates to physicians’ knowledge and attitudes, guideline-related factors and external factors.[3] A previous systematic review identified similar items with the additional element of patient barriers and classified them into seven categories, namely cognitive-behavioral barriers, attitudinal or rational-emotional barriers, professional barriers, barriers embedded in the guidelines or evidence, patient barriers, support or resources and system and process barriers.[9] These factors do not seem to vary much in the different areas of healthcare, be it general practice [5], long-term care [6] or for example prescribing [1].

Many different frameworks, theories or models have been developed to overcome these barriers and facilitate the translation process. Two recent reviews were carried out [10-11], both of which collected and classified these works according to Nilsen [12]. Huybrechts et al focused on the process models and the determinant frameworks, identifying their common elements. They found that the core phases of implementation are the development, translation and sustainment phase, while intended change, context and implementation strategies were highlighted as core components.[11] On the other hand, the aim of Esmail et al was to help users to select from the many existing concepts, so they categorized 36 works according to target audience, user level and Nilsen classification. Then comparison were made within each category to reveal similarities and uniqueness.[10] However, the situation is complicated by the fact that studies using implementation frameworks do not describe well their application and operationalization.[13-15] Reporting guidelines can alleviate the problem to some extent by helping readers assess the applicability of new knowledge to their own context.[16-18] We ourselves used SQUIRE 2.0 (Standards for Quality Improvement Reporting Excellence) when compiling our manuscript.[16]

Our study focused primarily on organizational implementation. However, we wanted to develop a method that would facilitate the implementation of an evidence in several institutions at the same time. We started from the assumption that although successful implementation means the new knowledge has to work in particular environments, it doesn’t mean that the whole translation process should exclusively be executed by the individual institutes or their representatives. Part of the process is still generalizable, either because the nature of the evidence allows it or because the context and actors show similarities. Accordingly, our aim was to develop a methodology that shows how to derive the general part of the implementation from the evidence. We named this general, intermediate state “general good practice”, which is – in our reading – a detailed frame for specific health service activities and systematic considerations of what and how to build on this frame. In this way, it can be clearly distinguished from the institutional good practice, which is usually referred to as good practice or best practice and which is the effective implementation of specific health care activities in a given institution. To get to general good practice, we first had to set up a framework that would organize the existing knowledge and our experience in implementation science in a way that would suitable for building such a methodology.

## Materials and Methods

We chose to use a disciplinary approach to gain the information on what knowledge is needed for a successful translation, because it was able to show what possible tools could be included in the development of general good practice (GGP). Glasziou et al explored the importance of the relationship between evidence-based medicine (EBM) and quality improvement (QI) pointing out that if EBM helps us “do the right things” while QI tells us to “do things right”, together we can “do the right things right”. [19] We examined in more detail the determinant frameworks that we considered most relevant to our context, since, according to Nilsen, they “specify types (also known as classes or domains) of determinants and individual determinants, which act as barriers and enablers (independent variables) that influence implementation outcomes (dependent variables). Some frameworks also specify relationships between some types of determinants. The overarching aim is to understand and/or explain influences on implementation outcomes, e.g. predicting outcomes or interpreting outcomes retrospectively.”[12] We have found that, alongside EBM and QI, change management (CM) is the main discipline with a broader perspective that includes e.g. organizational culture, leadership, project management, general and human resource management or behavioural science to be applied in implementation. To demonstrate, Table 1 shows how the elements of the different determinant frameworks relate to these three disciplines. We have listed the frameworks that were identified as determinant ones in the two reviews mentioned above[10-11] with the exception of five that were not found to be relevant, either because they lacked an organizational focus, included phases, levels or barriers rather than classical determinants, or were specific to social care [9, 20-23]. At the same time, two additional frameworks [24-25], previously known to us and considered relevant, were added, making a total of 16 conceptions examined [24-39].

**Table 1:**
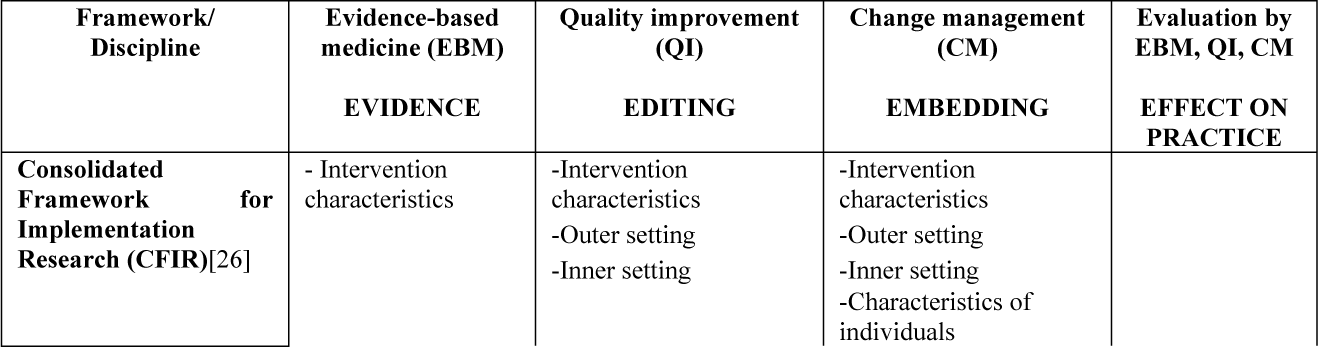
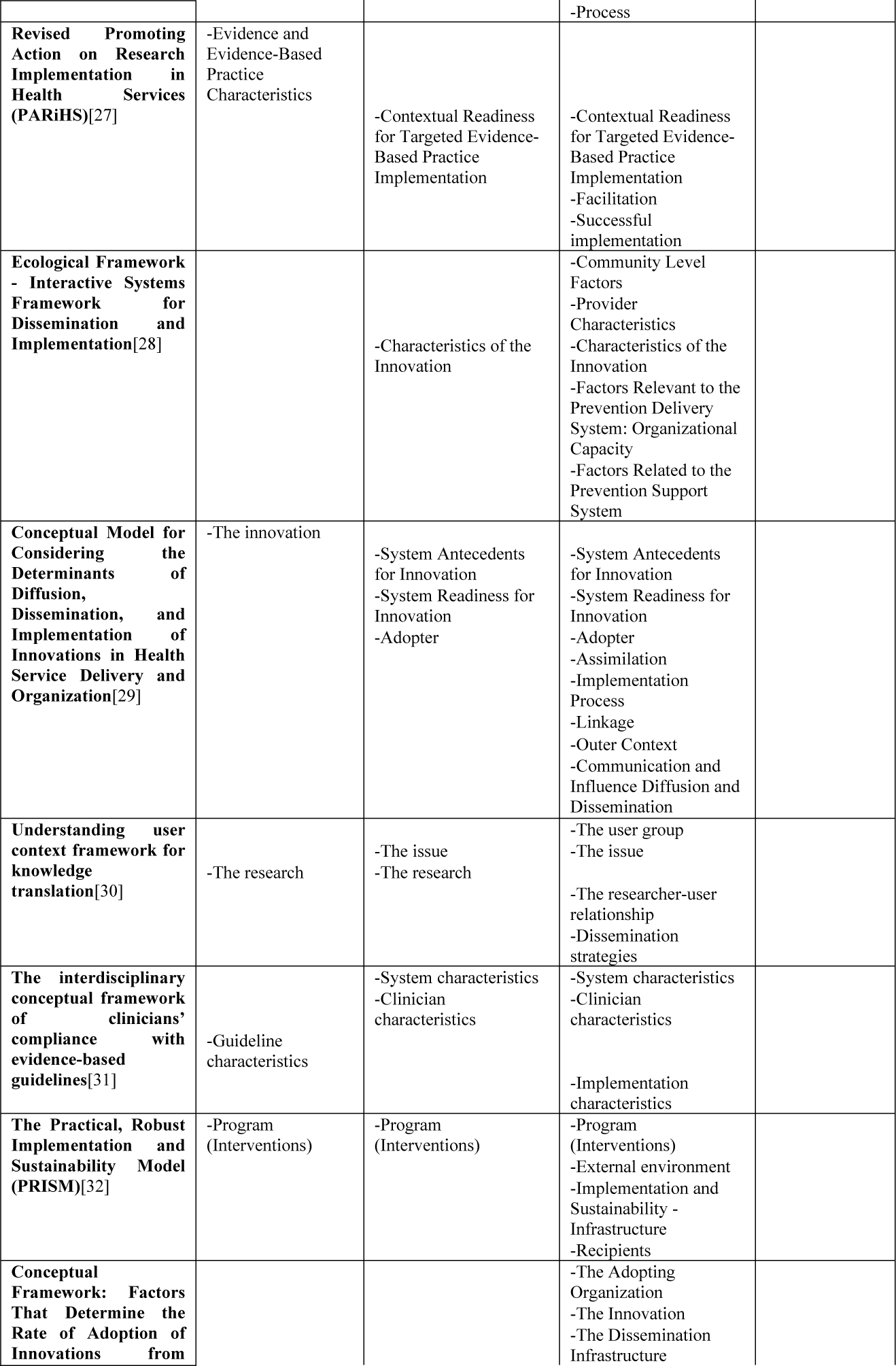
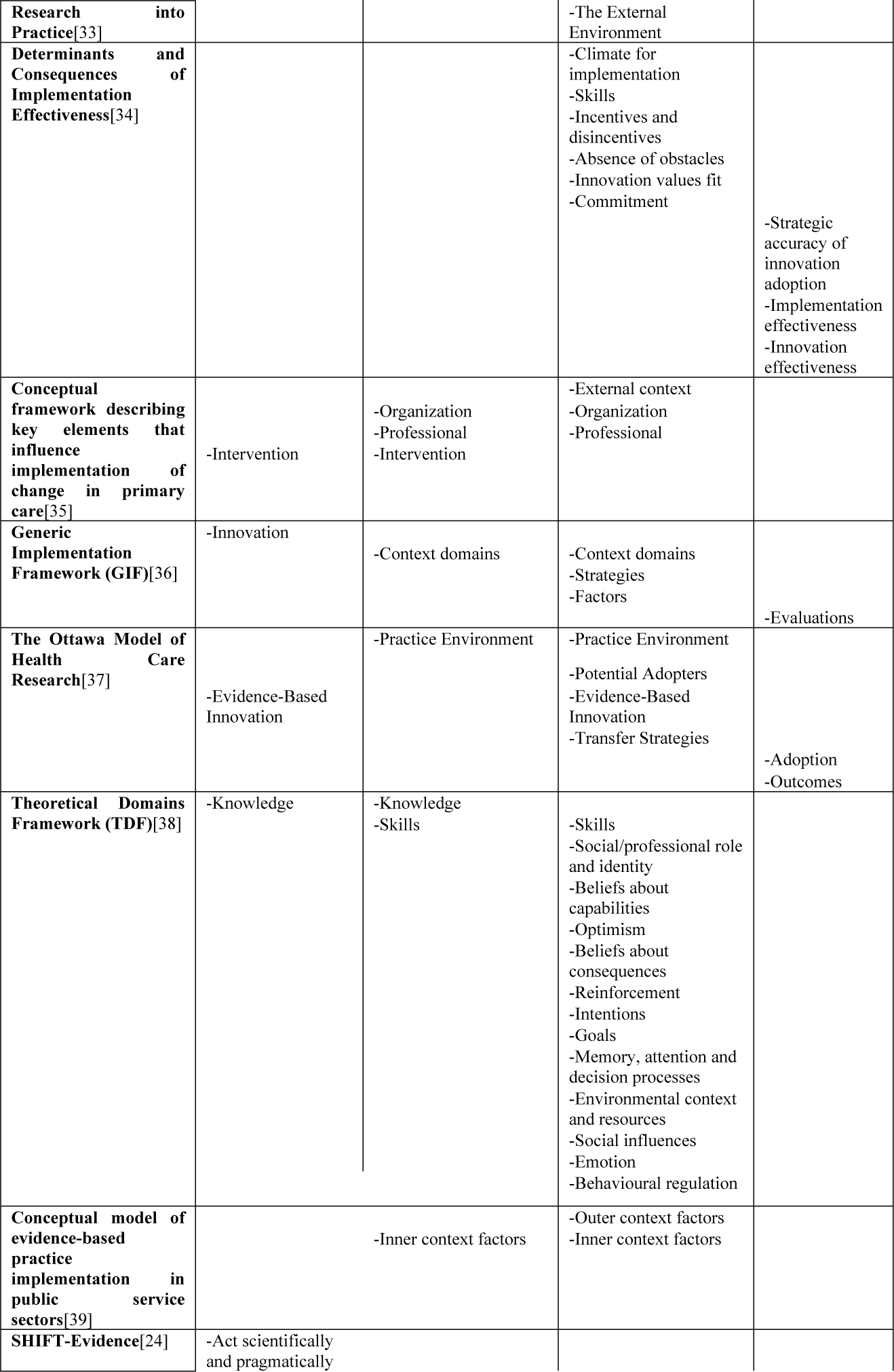
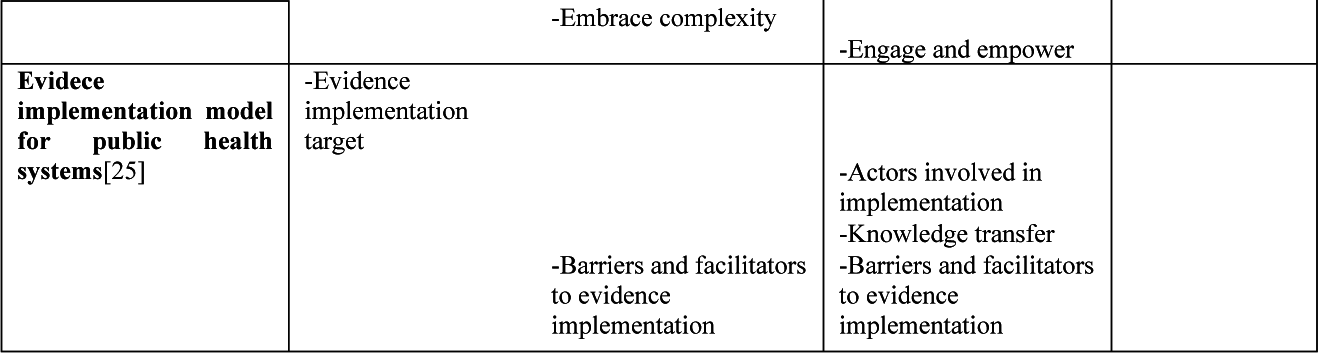
The elements of different determinant frameworks in the light of disciplines.

Once the *evidence* has been identified as worthy of implementation in the light of EBM, the elements belonging to QI allow us to tailor the practice so that it is capable to produce the evidence. This group of activities can therefore be called *editing*. However, at this point we are still standing at a theoretical station. In order for this to be translated into real practice, we need to change the existing practice accordingly. To express that this change must be permanent, we can use the term *embedding* to name this part. And this is precisely the area to which the elements of CM belong. Adding to EBM and QI, CM therefore can show us “to achieve right to do the right things right”. As a result, the *effect on practice* can be assessed using measurements of these three disciplines. As evidence, editing, embedding and their effect on practice are all connected to each other, exist simultaneously and forms an ever-recurring process, we represent them along an infinite sign, creating the concept of infinitE (Fig 1).

**Fig 1:**
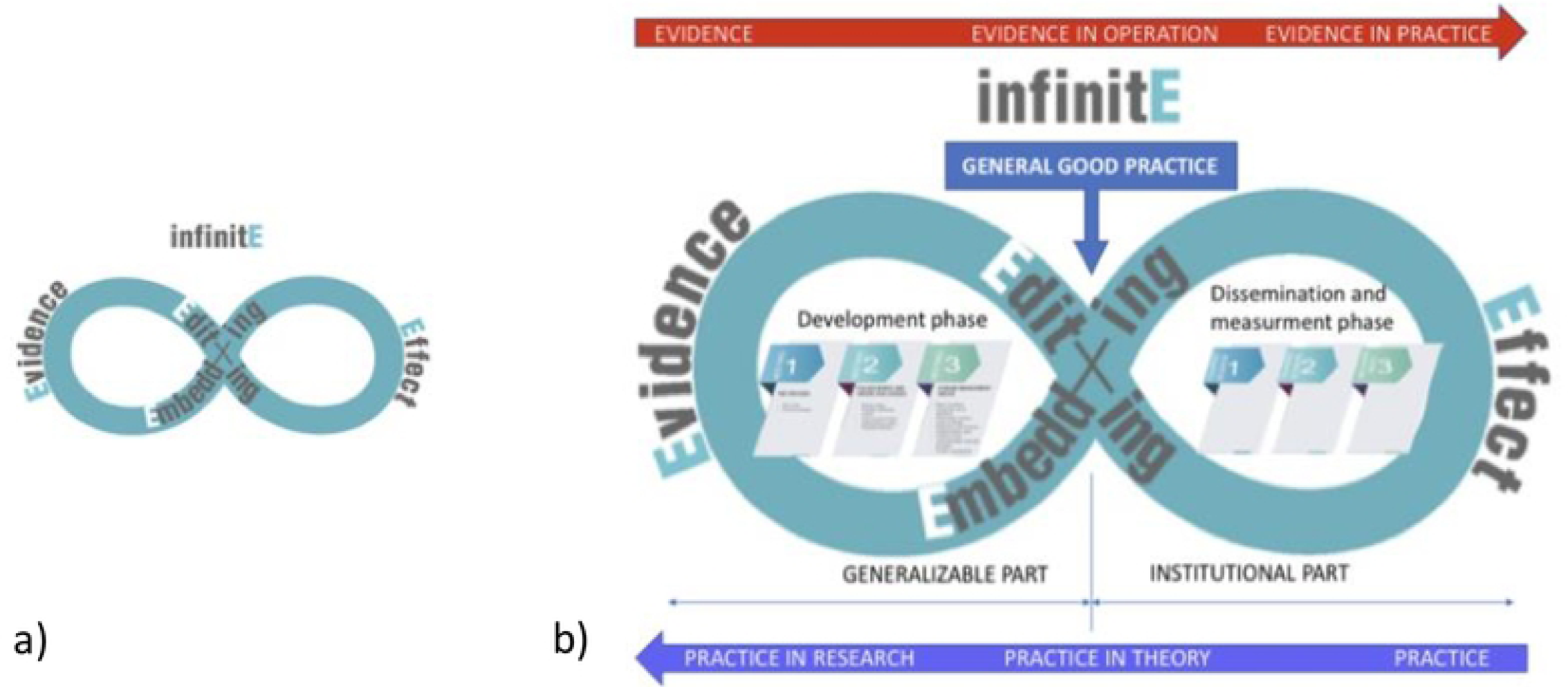
a) The concept of infinitE b) Research phases and general good practice in the light of the infinitE concept.

Based on this concept of ours, a methodology for the development of general good practice was developed in the framework of the European Union funded project “Professional Methodological Development of the Healthcare System” Patient Safety sub-project in Hungary. An initial methodology was put together by a *core group* of patient safety and quality management experts, and then validated by a *wider group of experts* from around the country with diverse healthcare experience, including professionals from all the four medical faculties in Hungary, with no proposal for change.

As the project’s expectations limited our scope somewhat, we drew *evidence* from two main sources. Firstly, we collected good practices from healthcare institutions through an online survey. In less than two months, 134 practices were submitted, all of which were assessed by two independent experts using an evaluation form, which was designed to map, among other things, the importance of the topic and the evidence behind it, the size of the patient population concerned, the range of specialties and occupational groups involved, the expected impact and the difficulties of design. The wider group of experts decided by consensus on which topic to develop further, considering the results of the evaluations. On the other hand, the guides produced in another strand of the sub-project were used as a source of evidence, as they were also expected to have associated good practices.

Regarding the *editing* part, we decided to first apply cause analysis in order to understand the factors that make the evidence not work well in practice and to respond to these by developing a detailed process of relevant care activities. To illustrate the process, we have proposed the ARIS business model diagram, which also facilitates process analysis by showing for each step the input and output event, the actors, and the input and output information or documentation needs.[40] The focus here was therefore on identifying those elements which, whatever the circumstances, seem to be generally necessary for the evidence to be take shape. As an additional aid, we have also designed a tabular representation of the information, where other elements of the process analysis not visible in the diagram, such as the devices, the location, the time or even the audit criteria, can be included. Next, we added the identification of possible failure modes to the previously identified process steps. Finally, according to the Donabedian model, a systematic definition of some structure, process and outcome indicators was placed at the end of the editing phase.

As for the *embedding* part, among the many change management frameworks, Kotter’s 8-step model was chosen for use, partly because it is sufficiently didactic to be followed by those less familiar with this discipline, and partly because the many areas and factors related to change management can be easily associated with the steps of the model, and thus provide a complex framework for potential users.[41] To set up a change management plan based on Kotter’s model, the following factors were considered:

- Basic conditions for implementation without which it is not worth starting
- Elements of corporate culture that are key to good practice
- Stakeholder analysis (potential stakeholders, their interests and influence)
- Level of change envisaged (e.g. individual, department, organizational)
- Possible forms of resistance and their possible solutions
- Proposed composition of the implementation team
- Associated training needs
- Potential communication channels and content (especially at the beginning, at the first success and on an ongoing basis)
- Further consideration for the 8-step model

The editing and embedding parts were designed to be carried out during a three months period with three face-to-face, full-day meetings, by a team with members from those who sent good practices in the related topic, experts in the field and moderator(s) with patient safety and quality management experience. In the period between the meetings, the preparation of related materials was done through a collaborative online editing interface.

The wider group of experts chose the institutional process of resuscitation as the topic for piloting the methodology, as it can affect every departments and professionals in a hospital, has a great emphasis on correct execution and on collaboration between actors, and because six different institutions submitted good practices in this area, including a cardiology institute, a mixed profile city hospital, two children’s hospitals, an ambulance service and an outpatient clinic, which provided a great opportunity to see how a process could be generalised. The three meetings of the pilot development were followed live from another room by the members of the expert groups, which not only allowed them to see the methodology in action, but also served as a model for future moderators, who were selected from the wider group of experts. After the pilot, the methodology of developing general good practice was finalized and applied to many other topics. We regularly discussed the experiences and comments of team members and expert groups in our meetings to draw conclusions and modify the development process where necessary.

The study of the *effect on practice* was part of a later stage of the research (Fig 1). It is because in order for the institutions to be able to use the results of the development of general good practices, their content and possible uses first had to be explained to them. To this end, training courses were designed and delivered, the methodology of which and the combined effect of the development and training of general good practices will be presented in a forthcoming article. For now, we focused our attention on the methodology for the development of general good practices.

## Results

The pilot development was successfully carried out with the planned three meetings in a three months period. The development team consisted of a delegated representative from each of the six institutions submitting a good practice, a moderator and an assistant moderator. The delegates also represented different occupational groups, including an anaesthesia and intensive care specialist - who was also a paediatrician-, a cardiologist, a director general, a neonatologist, an ER nurse, a healthcare manager – who was also a graduate nurse – and a quality officer – all of them played a key role in the development of their institution’s good practice. The moderator and the assistant moderator came from the core group of patient safety and quality management experts, and were in continuous contact with the rest of the core group.

The outputs of the general good practice development for the institutional process of resuscitation are shown in Table 2.

**Table 2:** Outputs of the pilot and the final proposed bundle of general good practice.

| OUTPUTS OF THE PILOT DEVELOPMENT | PROPOSED BUNDLE OF GENERAL GOOD PRACTICE |
| --- | --- |
| ▪ Detailed flow chart (S1 Fig.)<br>▪ Process table (S1 Table.)<br>▪ Table of the potential failure modes and the potential underlying causes (S2 Table.)<br>▪ Set of critical process steps and failure modes (focusing on the most likely to be significant elements on country level)<br>▪ Three indicator definitions (S3 Table.)<br>▪ Change management aspects and considerations (focusing on the most likely to be significant factors on country level)<br>▪ Text description of the general good practice (S1 File.)<br>▪ Instruction for use of the above materials | Core elements:<br>▪ Detailed flow chart (or list of system elements for a systems approach)<br>▪ Process table (or characterisation of system elements for a systems approach)<br>▪ Table of the potential failure modes and the potential underlying causes<br>▪ Change management aspects and considerations (from a general, e.g. country-level or profession-specific perspective)<br>Additional elements:<br>▪ Indicator definitions<br>▪ Text description of the general good practice<br>▪ Instruction for use of the above materials (not necessarily topic-specific)<br>▪ Set of critical process steps and failure modes (from a general, e.g. country-level perspective) |

The pilot project resulted in three changes to the development methodology. Flow chart and process analysis seemed to be the primary steps to be applied, while the possible underlying causes seemed to be more reasonably attributed to the already identified failure modes. Failure modes were attached to each process step, but it seemed unnecessary to count the possible underlying causes for each failure mode because there was too much repetition. Rather it was reasonable to identify them as a group belonging to the failure modes of a particular process step. The last change was of a technical nature: instead of a whiteboard and flipchart, we used a digital solution, taking notes on a laptop, which could be simultaneously viewed and validated by the participants via a projector. Accordingly, templates were prepared to facilitate and standardise the steps of development. The final development process is illustrated in Fig 2.

**Fig 2:**
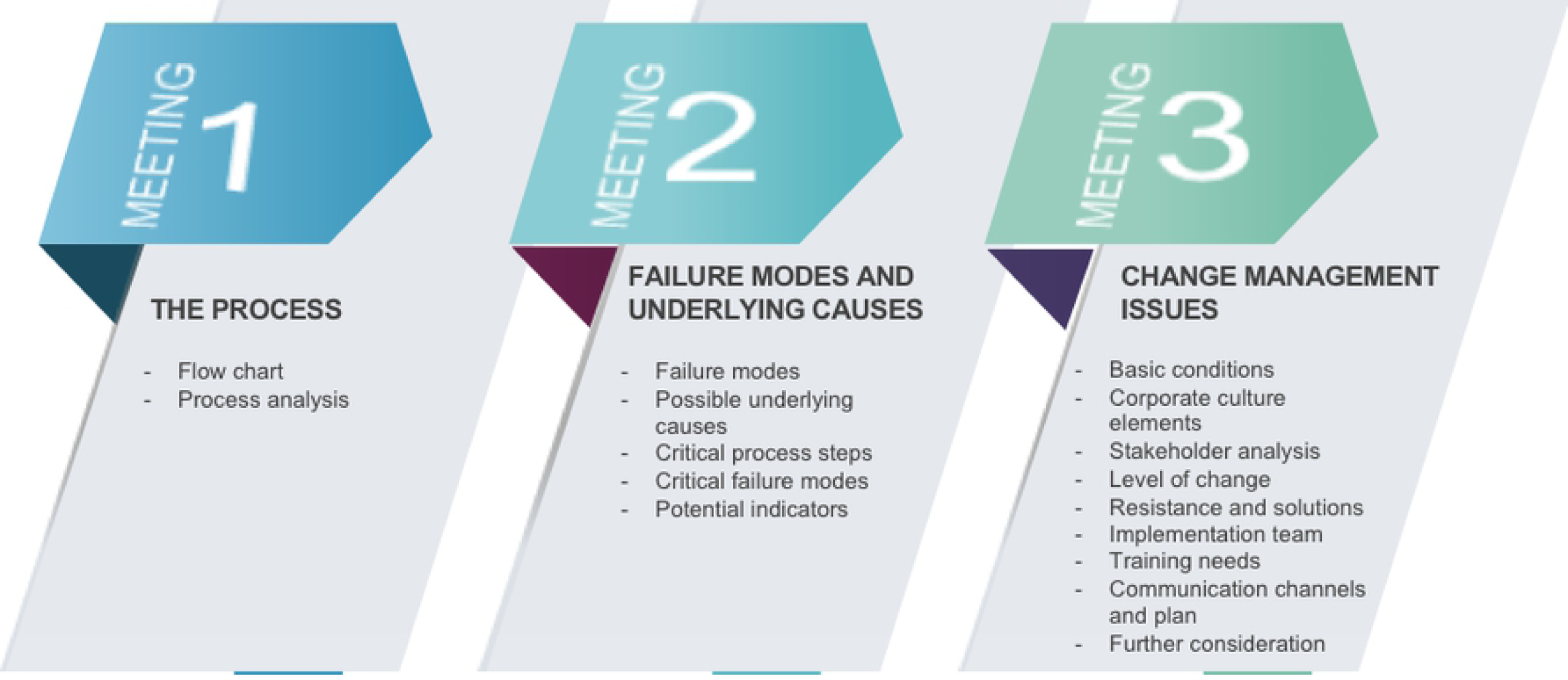
Developing general good practices: the editing and embedding part.

After the pilot, the development methodology was applied to more than ten other topics, including pressure ulcer prevention and care, perioperative pain management, two-step onco-team practice, patient education, inpatient hand hygiene, personalized medication or some prevention processes for various hospital-acquired infections. These allowed further conclusions to be drawn. First of all, not all the topics could be approached from a process perspective. Patient education and hand hygiene seemed to be better processed from a systems-approach. In these cases, the flow chart and process analysis have been replaced by the identification and detailed study of system elements. Secondly, and unfortunately, the systematic definition of indicators seemed to be an explicitly advanced area as in most cases even the good-practice institutes did not apply such monitoring activities, and if they had, the way to standardise measurements was so elusive that it seemed very far from being possible to define a formula that could be generally applied across institutions. Therefore, in the majority of themes, the systematic development of indicators was ultimately abandoned, and only a list of names of potential indicators was drawn up. Finally, it became evident that even in cases where the developers from the institutions included people with quality experience, the moderators played a crucial role in ensuring that the use of the various QI tools was properly understood and applied. Based on this experience we have finally defined general good practice as a bundle of core elements that can always be derived from development, providing essential content and which can be supplemented with additional considerations (Table 2).

## Discussion

The use of ARIS process modelling was found to be appropriate in several respects. It is suitable for showing the temporality of the process from the initial event downwards, together with the steps that can be carried out in parallel. Logical links between steps (and, or, or else) can also be detected and alternative paths can be followed. The process table structures information in a way that allows to examine a particular step in the process in detail (looking at a given row), or to monitor a type of data, like actors or required information, throughout the process (focusing on one column). We found that the flow chart and the process table can be used in many ways as shown in Table 3.

**Table 3:** Possible uses of the flow chart and the process table.

| Possible uses of the flow chart and the process table |  |
| --- | --- |
| Complete in-hospital development of the given care process | If the healthcare provider has not previously operated the care process in question, the flow chart and process table will help to guide the person(s) responsible for the implementation through all process steps from start to finish. |
| Assessing, streamlining or improving the implementation of a given care process in the institution | If the care process in question is already in some form of operation, comparing the current process step by step with the flow chart and process table, we can identify the gaps that to improve the existing process. In doing so, it is worth reviewing whether the own current process includes all the steps set out in general good practice, contains steps that are missing from the general good practice and can be omitted or has the temporal and logical links between its steps as outlined in the general good practice. |
| Defining roles, responsibilities and competences, defining job descriptions | Whether you are setting up a new process or improving an existing one, thinking about the actors involved in the process steps will help ensure, for example, that no process step is left without a responsible person, and responsibilities are transparent where there are several possible actors. |
| Reviewing documentation requirements, ensuring the availability and development of the necessary knowledge | Whether we are developing a new process or improving an existing one, reviewing the knowledge, required or generated documents for each process step will help, for example,<br>▪ identify the documents that contain the necessary knowledge, whether they are educational materials, protocols or items in the patient's medical record, and make them available,<br>▪ ensure the availability of the documentation required for each step,<br>▪ identify whether the implementation of the process step requires documentation and, if so, assign the required content, format and person (job group) responsible for the documentation,<br>▪ coordinate the activities of the persons responsible for the process step and the documentation of the process step, by providing documentation rights and access. |
| Organisation and development of training | After an overview of the process, the actors, the necessary knowledge and the documentation requirements, the content of the related training can be identified by job group (actor). |
| Design and development of the care process monitoring system | The information in the flow chart and process table can be used to derive the structural characteristics of the care in question. For human resources, the overview of actors can provide information, while for physical assets and conditions, the identification of devices and locations can be used as a source. As for regulation, the necessary knowledge (e.g. procedures, protocols) and documentation requirements (e.g. document templates, samples) can provide a basis for monitoring. In order to define process indicators, the process steps for which measurability is theoretically meaningful can be identified. The nature of the output events, as well as the timeliness, documentation and characteristics of the persons responsible, can form the basis for demonstrating compliance. As expected, the identification of outcome indicators is the most difficult, as many factors other than the process of care are involved in the change in the patient's health status. Yet, looking at the impact of individual steps on outcomes can help to do this. This may include considering how the correct implementation of a particular process step can avoid adverse events or add value to the patient's recovery. |

Comparing a given institutional practice with a flowchart and process table can give us an answer to whether that practice can provide the right care. This approach can be complemented by an enumeration of failure modes, which in turn will answer whether the institutional practice allows for the avoidance of failures. The failure modes collected in general good practice aim to cover the theoretically possible failure modes, so that we can review them to assess which ones are relevant in a given institutional practice and how important they are. The underlying causes associated with failure modes are more of a food for thought, but if a failure mode is found to be significant in institutional practice, a detailed root-cause analysis will be needed to find the right local solution.

Proposals based on change management knowledge to support the implementation of good practice provide a menu for potential users to identify the elements that need to be addressed in their institution and to select a combination of options and approaches to address them.

As mentioned earlier, the proper application of quality improvement tools and the professionalism of the products produced required the intensive involvement of moderators, even when the developers included people with quality experience. Yet the most unknown and innovative element was undoubtedly the area of change management, and this is also true for quality professionals.

To formulate how the general good practice differs from or adds to the guideline, it is perhaps easiest to say that while the guideline formulates the evidence, general good practice shows the evidence in operation. On the pilot topic of resuscitation, for example, the 2020 American Heart Association guidelines for cardiopulmonary resuscitation and emergency cardiovascular care present the process, recommendations and knowledge gaps that can be translated into practice, but say the evaluation of their feasibility and acceptability is not in their scope.[42] Similarly, the European Resuscitation Council (ERC) Guidelines for Resuscitation 2015 state that “the combination of medical science and educational efficiency is not sufficient to improve survival if there is poor or absent implementation”, but mention only a few, mainly systemic points in this context such as trainings in schools or establishing cardiac arrest centres.[43] The new version 2021 already includes some more concrete considerations for the institutional implementation in terms of first responder, equipment and the resuscitation team, but it remains an open question for those doing the translation on how best to design these elements in their own institution.[44] As an example, for one topic, Table 4 shows the difference between the latter, the most advanced guideline in this respect, and the general good practice.

**Table 4:** An example of the difference between guidelines and general good practices in the pilot topic of resuscitation.

| WHAT THE GUIDELINE SAYS* | TOPIC IN QUESTION | WHAT THE GENERAL GOOD PRACTICE HELPS TO THINK THROUGH |
| --- | --- | --- |
| It may differ between hospitals or locations within a hospital<br>If the responder is alone, they may need to leave the patient<br>Where a telephone system is used, the standard European number should be used | CALL FOR HELP | What ensures that the first responder knows the alarm channel?<br>What ensures that the first responder knows when to call for help?<br>What if the first responder is not a health worker (e.g. visitor, cleaning lady, another patient, etc.)?<br>Is the alarm device accessible everywhere or is the location of the nearest one known (even in places such as parks or canteen, etc.)?<br>What ensures that the alarm number is known by everyone?<br>Is the alarm channel one-way so that it cannot be occupied?<br>What ensures that the alarm device is accessible for the receiver at any moment?<br>What ensures that the alarm device is always operational on both sides (e.g. maintained, charged, volume is on, adequate network coverage, signal strength, etc.)?<br>Is there any difference if the first responder is alone or with someone else?<br>What ensures that the first responder knows what to say and how to say it during an emergency call, so that they can give the necessary and correct information as quickly as possible?<br>What ensures that the first responder can do this properly at any time, even in real, stressful situation? |

\*These findings are taken from the European Resuscitation Council Guidelines 2021: Adult advanced life support[44]

From the above, it seems that there is indeed a generalisable part of the implementation process, and the general good practice is a good representation of this. Accordingly, in the infinitE model, it can be fitted to the half-way point of the process, symbolically separating the generalisable and institutional parts of the translation. Furthermore, it shows the evidence in operation, which will then be put into practice by the editing and embedding processes of the institute. The venous system of the same mechanism will ensure that the practice is incorporated into theoretical considerations, while the evolution of general good practice can be embodied in the directions, elements and design of further researches (Fig 1).

In our view, the novelty of the infinitE model presented in our paper lies in the fact that it presents the elements of translation from a focus on creating practical applicability in a simple and pragmatic way, successfully marrying CM with the EBM-QI dual already paired before.[19] This is also reflected in the general good practice developed on the basis of the model, as its methodology successfully combines the three disciplines. Thanks to this, the methodology was applicable to several other topics, thus the core elements of the general good practice bundle were always produced as a result of the development. Also, the methodology integrates all the known factors from the related literature introduced earlier. As the use of general good practices in different institutions can be paralleled with practice development, its relation to it may be interesting. We can see that the formula also fits in well with the recommendations of practice development, for example, it is suitable for the joint dissemination of process and product knowledge[8], it takes cultural aspects into account[45] and can also serve the main characteristics of practice development as presented by Page[46]. However, in addition to these, our work also defines a significant additional step in the translation process, which is, to our knowledge, the first attempt to do so.

Perhaps, the biggest limitation of our research was that we conducted the pilot and the subsequent general good practice developments in a country with limited resources for health care and with persistent and substantial human resource problems.[47-50] Furthermore, the private sector was not involved in the study as the participants of the development teams were all employees of public healthcare institutions. Therefore, the outcomes of the developments may not applicable to other health systems without any corrections. It is conceivable that, for example, the layout of the processes involved could be modified by different technological backgrounds. Even in our case, two versions of the general good practice of personalized medication were produced, depending on whether it was manual or automated medication. Also, the number of professionals available and their different qualifications can affect the division of labour and the level of decision-making. On the other hand, in the case of more advanced quality system and experience, the general good practices can become even more complete, for example with developing specific indicators or even monitoring systems as well as patient registers or standardised documentation. We have only been able to do the latter in one case, perioperative analgesia, which, although it meant extra time, could contribute to improving the poor situation of Acute Pain Service in Hungary.[51] These considerations lead to conclusion that general good practices should be developed or adapted at regional or national level, or specific to a health system, but the development methodology itself is likely to be generally applicable.

## Conclusions

The concept of general good practice was found to be sound, and the development methodology was seen to be applicable to a wide range of topics. General good practice represents a new, unprecedented step in the translation process that can make it easier for the institute’s quality and patient safety staff, as well as the chief medical officers and head nurses, to put professional innovations into practice, whether it is the introduction of a new guideline or best practice, or the introduction of a new technology or device. In addition, however, it can contribute to the definition of possible process indicators of care, and thus to its monitoring, as well as to the development of documentation, including standardised documentation. Such systematic mapping of processes can also be a starting point for the digitalisation of care processes. The question arises as to who should be responsible for developing general good practices. There are different options: guideline developers may do it as a final step in the development process, but it can also be the responsibility of medical universities, operator of healthcare institutions or health care workers’ professional organisations. The involvement of Research Translation Centres may also be an obvious solution, as they were set up to accelerate the translation of evidence by creating partnerships between research institutes, universities and health services.[52] Whichever path we choose, it is important to ensure that the development team represents the knowledge and skills of EBM, QI, the related practice and CM.

In our next step, we have designed a training methodology to familiarise healthcare institutions with general good practice and how they can use it, thus, how they can base their own editing and embedding activities on it, in order to better reflect the evidence in their care (Fig 1). In agreement with Burke et al, while investigating the effects on practice, we focused on sustainable implementations, that remain effective for at least six months.[53] Other studies could investigate the applicability of the development methodology to other topics, especially in the case of a systems approach, as there were few opportunities to do so to date.

## Data Availability

Most relevant data are within the manuscript and its Supporting Information files. However, sources such as minutes, notes, working documents are available in Hungarian, contain sensitive data, can be confidential and are also part of the project administration. Requests for data access can made in writing to Semmelweis University Health Services Management Training Centre. Requests will be considered, and de-identified datasets will be prepared and made available electronically in Hungarian.

## Acknowledgements

The authors would like to thank all the members of the core and wider group of experts in the study for providing input at any stage of the development. We are also grateful to all the health professionals who participated in the development of general good practices on the different topics. Moreover, we wish to thank all senior colleagues in our institute for their general support in the conduct of our study.

## Supporting information

**S1 File. Flow chart of the institutional process of resuscitation** (PDF)

**S1 Table. Process table of the institutional process of resuscitation** (PDF)

**S2 Table. Table of the potential failure modes and underlying causes of the institutional process of resuscitation** (PDF)

**S3 Table. Indicators for the institutional process of resuscitation** (PDF)

**S2 File. Text description of the general good practice of the institutional process of resuscitation** (PDF)

**S3 File. Good practice submission form** (PDF)

**S4 File. Evaluating sheet for the evaluation of submitted good practices** (PDF)

## Notes

### Competing Interest Statement

I have read the journal’s policy and the authors of this manuscript have the following competing interest: Ã.B. is the President and J.L. is the Vice President of NEVES EgyesÃ1/4let, which is a non-profit patient safety organization in Hungary.

### Funding Statement

Our research was carried out under the European Commission-funded EFOP-1.8.0-VEKOP-17-2017-00001, “Professional Methodological Development of the Healthcare System” project. All authors (HS, JL, IB, éB) and activities regarding the development and the related meetings were funded by this programme. The funder had no role in study design, data collection and analysis, decision to publish, or preparation of the manuscript.

### Author Declarations

Not relevant, as our institution does not require ethical approval if patients are not involved in the research.

## References

1. Paksaite P, Crosskey J, Sula E, West C, Watson M. A systematic review using the Theoretical Domains Framework to identify barriers and facilitators to the adoption of prescribing guidelines. Int J Pharm Pract. 2020.

2. Jin YH, Tan LM, Khan KS, Deng T, Huang C, Han F et al. Determinants of successful guideline implementation: a national cross-sectional survey. BMC Med Inform Decis Mak. 2021;21(1):19.

3. Fischer F, Lange K, Klose K, Greiner W, Kraemer A. Barriers and Strategies in Guideline Implementation - A Scoping Review. Healthcare. 2016;4(3):36.

4. Rosa RG, Teixeira C, Sjoding M. Novel approaches to facilitate the implementation of guidelines in the ICU. J Crit Care. 2020; 60:1–5.

5. Grol R. Implementing guidelines in general practice care. Qual in Health Care. 1992;1(3):184–191.

6. McArthur C, Bai Y, Hewstone P, Giangregorio L, Straus S, Papaioannou A. Barriers and facilitators to implementing evidence-based guidelines in long-term care: a qualitative evidence synthesis. Implement Sci. 2021;16(1):70.

7. Graham ID, Logan J, Harrison MB, Straus SE, Tetroe J, Caswell W, et al. Lost in Knowledge Translation: Time for a Map? J Contin Educ Health Prof. 2006;26(1):13–24.

8. Newell S, Edelman L, Scarbrough H, Swan J, Bresnen M. ‘Best practice’ development and transfer in the NHS: the importance of process as well as product knowledge. Health Serv Manage Res. 2003;16(1):1–12

9. Cochrane LJ, Olson CA, Murray S, Dupuis M, Tooman T, Hayes S. Gaps Between Knowing and Doing: Understanding and Assessing the Barriers to Optimal Health Care. J Contin Educ Health Prof. 2007;27(2):94–102

10. Esmail R, Hanson HM, Holroyd-Leduc J, Brown S, Strifler L, Straus SE, et al. A scoping review of full-spectrum knowledge translation theories, models, and frameworks. Implement Sci. 2020;15(1):11

11. Huybrechts I, Declercq A, Verté E, Raeymaeckers P, Anthierens S. The Building Blocks of Implementation Frameworks and Models in Primary Care: A Narrative Review. Front Public Health. 2021;9:675171

12. Nilsen P. Making sense of implementation theories, models and frameworks. Implement Sci. 2015;10:53

13. Hull L, Goulding L, Khadjesari Z, Davis R, Healey A, Bakolis I, et al. Designing high-quality implementation research: development, application, feasibility and preliminary evaluation of the implementation science research development (ImpRes) tool and guide. Implement Sci. 2019;14(1):80.

14. Bergström A, Ehrenberg A, Eldh AC, Graham ID, Gustafsson K, Harvey G, et al. The use of the PARIHS framework in implementation research and practice – a citation analysis of the literature. Implement Sci. 2020;15(1):68.

15. Moullin JC, Dickson KS, Stadnick NA, Rabin B, Aarons GA. Systematic review of the Exploration, Preparation, Implementation, Sustainment (EPIS) framework. Implement Sci. 2019;14(1):1

16. Ogrinc G, Davies L, Goodman D, Batalden P, Davidoff F, Stevens D. SQUIRE 2.0 (Standards for QUality Improvement Reporting Excellence): revised publication guidelines from a detailed consensus process. BMJ Qual Saf. 2016;25(12):986–992.

17. Brouwers MC, Kerkvliet K, Spithoff K. AGREE Next Steps Consortium. The AGREE Reporting Checklist: a tool to improve reporting of clinical practice guidelines. BMJ. 2016;352:1152.

18. Chen Y, Yang K, Marusic A, Qaseem A, Meerpohl JJ, Flottorp S, et al. RIGHT (Reporting Items for Practice Guidelines in Healthcare) Working Group. A Reporting Tool for Practice Guidelines in Health Care: The RIGHT Statement. Ann Intern Med. 2017;166(2):128–132.

19. Glasziou P, Ogrinc G, Goodman S. Can evidence-based medicine and clinical quality improvement learn from each other? BMJ Qual Saf. 2011;20 Suppl 1(Suppl_1):13–17.

20. Social Marketing National Excellence Collaborative. Social Marketing and Public Health. Lessons from the Field. A Guide to Social Marketing. Turning Point National Program Office at the University of Washington; 2003. Available from: https://www.dors.it/marketing_sociale/docum/2_smc_lessons_from_field.pdf

21. Glasgow RE, Vinson C, Chambers D, Khoury MJ, Kaplan RM, Hunter C. National Institutes of Health Approaches to Dissemination and Implementation Science: Current and Future Directions Am J Public Health. 2012;102(7):1274–1281

22. Blom B, Morén S. Explaining Social Work Practice - the CAIMeR Theory. J Soc Work. (2010) 10(1):98–119

23. Ferlie EB, Shortell SM. Improving the quality of health care in the United Kingdom and the United States: a framework for change. Milbank Q. 2001;79(2):281–315

24. Reed JE, Howe C, Doyle C, Bell D. Successful Healthcare Improvements From Translating Evidence in complex systems (SHIFT-Evidence): simple rules to guide practice and research. Int J Qual Health Care. 2019;31(3):238–244.

25. Vincenten J, MacKay JM, Schröder-Bäck P, Schloemer T, Brand H. Factors Influencing Implementation of Evidence-Based Interventions in Public Health Systems – A Model. Centr Eur J Public Health. 2019;27(3):198–203

26. Damschroder LJ, Aron DC, Keith RE, Kirsh SR, Alexander JA, Lowery JC. Fostering implementation of health services research findings into practice: a consolidated framework for advancing implementation science. Implement Sci. 2009;4:50.

27. Stetler CB, Damschroder LJ, Helfrich CD, Hagedorn HJ. A Guide for applying a revised version of the PARIHS framework for implementation. Implement Sci. 2011;6:99.

28. Durlak JA, DuPre EP. Implementation matters: a review of research on the influence of implementation on program outcomes and the factors affecting implementation. Am J Community Psychol. 2008;41(3-4):327–50.

29. Greenhalgh T, Robert G, Macfarlane F, Bate P, Kyriakidou O. Diffusion Of Innovations In Service Organizations: Systematic Review And Recommendations. Milbank Q. 2004;82(4):581–629.

30. Jacobson N, Butterill D, Goering P. Development of a framework for knowledge translation: understanding user context. J Health Serv Res Policy. 2003;8(2):94–9.

31. Gurses AP, Marsteller JA, Ozok AA, Xiao Y, Owens S, Pronovost PJ. Using an interdisciplinary approach to identify factors that affect clinicians’ compliance with evidence-based guidelines. Crit Care Med. 2010;38(8 Suppl):S282–91.

32. Feldstein AC, Glasgow RE. A practical, robust implementation and sustainability model (PRISM) for integrating research findings into practice. Jt Comm J Qual Patient Saf. 2008;34(4):228–43.

33. Bradley EH, Webster TR, Baker D, Schlesinger M, Inouye SK, Barth MC, et al. Translating research into practice: speeding the adoption of innovative health care programs. Issue Brief (Commonw Fund). 2004;(724):1–12.

34. Klein KJ, Sorra JS. The challenge of innovation implementation. Acad Manage Rev. 1996;21(4):1055–1080

35. Lau R, Stevenson F, Ong BN, Dziedzic K, Treweek S, Eldridge S, et al. Achieving change in primary care--causes of the evidence to practice gap: systematic reviews of reviews. Implement Sci. 2016;11:40.

36. Moullin JC, Sabater-Hernández D, Fernandez-Llimos F, Benrimoj SI. A systematic review of implementation frameworks of innovations in healthcare and resulting generic implementation framework. Health Res Policy Syst. 2015;13:16.

37. Logan J, Graham ID. Toward a comprehensive interdisciplinary model of health care research use. Sci Commun. 1998;20:227–46.

38. Atkins L, Francis J, Islam R, O’Connor D, Patey A, Ivers N, et al. A guide to using the Theoretical Domains Framework of behaviour change to investigate implementation problems. Implement Sci. 2017;12(1):77.

39. Aarons GA, Hurlburt M, Horwitz SM. Advancing a conceptual model of evidence-based practice implementation in public service sectors. Adm Policy Ment Health. 2011;38(1):4–23.

40. Davis R. Business Process Modelling with Aris: A Practical Guide. London: Springer-Verlag London Ltd.; 2001.

41. Kotter JP. Leading Change: Why Transformation Efforts Fail. Harv Bus Rev 1995;May-June.

42. Merchant RM, Topjian AA, Panchal AR, Cheng A, Aziz K, Berg KM, et al. Part 1: Executive Summary: 2020 American Heart Association Guidelines for Cardiopulmonary Resuscitation and Emergency Cardiovascular Care. Circulation. 2020;142(16_suppl_2):S337–S357.

43. Greif R, Lockey AS, Conaghan P, Lippert A, De Vries W, Monsieurs KG, et al. European Resuscitation Council Guidelines for Resuscitation 2015: Section 10. Education and implementation of resuscitation. Resuscitation. 2015;95:288–301.

44. Soar J, Böttiger BW, Carli P, Couper K, Deakin CD, Djärv T, et al. European Resuscitation Council Guidelines 2021: Adult advanced life support. Resuscitation. 2021;161:115–151.

45. Cioffi J, Leckie C, Tweedie J. Practice development: a critique of the process to redesign an assessment. Aust J Adv Nurs. 2007;25(2).

46. Page S. The role of practice development in modernising the NHS. Nurs Times. 2002;98(11):34.

47. Gaál P, Szigeti Sz, Csere M, Gaskins M, Pantelli D. Hungary health systems review. Health Syst Transit. 2011;13(5):1–266.

48. Gaál P, Szigeti Sz, Panteli D, Gaskins M, van Ginneken E. Major challanges ahead for Hungarian healthcare. BMJ. 2011;343(7836):1251–1254.

49. Girasek E, Szócska M, Kovács E, Gaál P. The role of controllable lifestyle in the choice of specialisation among Hungarian medical doctors. BMC Med Educ. 2017;17(1).

50. Gaál P, Velkey Z, Szerencsés V, Webb E. The 2020 reform of the employment status of Hungarian health workers: Will it eliminate informal payments and separate the public and private sectors from each other? Health Policy. 2021;125(7):833–840.

51. Lovasi O, Lám J, Schutzmann R, Gaál P. Acute Pain Service in Hungarian hospitals. PLoS One. 2021;16(9).

52. Robinson T, Bailey C, Morris H, Burns P, Melder A, Croft C, et al. Bridging the research-practice gap in healthcare: a rapid review of research translation centres in England and Australia. Health Res Policy Syst. 2020;18(1):117

53. Burke RE, Marang-van de Mheen PJ. Sustaining quality improvement efforts: emerging principles and practice. BMJ Qual Saf. 2021;30:848–852.

